# SHP2 promotes NETosis through ERK5 pathway and mediates the development of psoriasis

**DOI:** 10.1101/2021.11.10.21266198

**Authors:** Yan Ding, Jiao Qu, Chenyang Zhang, Yuyu Zhu, Qiang Xu, Yang Sun

**Author notes:** Corresponding to Yang Sun, Ph.D., Professor, School of Life Sciences, Nanjing University, Nanjing 210023 China. These authors contributed equally to this work (Yan Ding, Jiao Qu).

## Abstract

Psoriasis is a chronic inflammatory disease which infiltrated a large number of neutrophils among skin lesions. Here, we investigated the contribution of tyrosine phosphatase SHP2 in neutrophils, as well as it’s pathogenesis in psoriasis. We combined single-cell RNA sequencing with experimental verification to declare that SHP2 in neutrophils could promote the NETs formation through the ERK5 pathway, and resulted in the infiltration of inflammatory immune cells, which leads to psoriasis. Our study provides evidence for the role of SHP2 in NETosis in the progression of psoriasis, and SHP2 may be a potential therapeutic target for the treatment of psoriasis.

## Introduction

Psoriasis is a common chronic relapsing inflammatory skin disease that is currently defined as an autoimmune disease. Psoriatic lesions are clinically characterized by red plaques covered with silvery-white scale and circumscribed papules. Patients usually present with itching and swelling. The histopathological features of psoriasis are hyperkeratosis of keratinocytes in the epidermal base, dilated and elongated capillaries in dermal layer, and infiltrated of inflammatory cells in skin lesion area. Traditional medication for psoriasis includes acitretin, methotrexate, and cyclosporine. In recent years, some biologics, such as: etanercept, infliximab have been used clinically. However, due to relapse after discontinuation of drugs, as well as the toxic side effects of drugs, the long-term use of the drugs is restricted, resulting in that psoriasis cannot be cured at present.The pathogenesis of psoriasis is still inconclusive. In recent years, studies have shown that abnormal immune system is a characteristic of psoriasis skin lesions, and it is also the main entry point for research on the pathogenesis of psoriasis. Neutrophils, as the most abundant cells in innate immune cells, also play an important role in the occurrence and development of psoriasis.

The traditional defense mechanisms of neutrophils include pathogen phagocytosis, degranulation, and cytokine production. In recent years, studies have found that neutrophils can undergo a special cell death process called “NETosis” when they are stimulated by inflammatory factors, chemicals, or metabolites.

In this process, PAD4 is responsible for the citrullination of histones in neutrophils, and histone 3 (H3) in the nucleus is modified to form citrullinated histone 3 (Cit-H3), which in turn causes the chromatin to become loose. Subsequently, myeloperoxidase (MPO) and neutrophil elastase (NE), which enter the nucleus to cleave histones to separate them from DNA, cause further chromatin decondensation[1]. At last, DNA releases to the outside of the cell[2]. At the same time, the granular proteins in the cytoplasm, including enzymes and antibacterial peptides, attach to the scaffold composed of chromatin to form a network-like complex called NETs. Studies have shown that NETs play an important role in the pathogenesis of psoriasis.

SHP2 (Src homology phosphotyrosine phosphatase 2) is a non-transmembrane protein tyrosine phosphatase, which is encoded by the human *PTPN11* gene, and consists of two N-terminal SH2 domains, a C-terminal tail and a catalytic protein tyrosine phosphatase (PTP) domain. SHP2 is involved in cell proliferation, differentiation and survival. More and more evidences show that SHP2 is also involved in the regulation of immune cells and the occurrence of inflammation. For example, SHP2 positively regulates the oxidative burst of macrophages[3] and promotes the migration of dendritic cells to secondary lymphoid organs[4]. SHP2 can also bind to multiple receptors related to immune response to activate T cells, and participate in the downstream signal transduction of PD-1[5].

Mitogen activated protein kinase (MAPK) signaling pathway is one of the important signal transduction systems in organisms. Its subfamilies mainly include extracellular signal regulated kinase 1/2 (ERK1/2), c-jun amino terminal kinase (JNK), p38 mitogen activated protein kinase (p38 MAPK) and extracellular signal regulated kinase (ERK5). MAPK family can participate in cell proliferation, differentiation, apoptosis and other functions. ERK5 is encoded by *MAPK7* in human, and it is located on 17p11.2 human chromosome and consists of 816 amino acids. ERK5 is expressed in most human cells and can be activated by mechanical stimulation, oxidative stress, hypertonic environment, cytokines and inflammatory cytokines (such as IL-6). ERK5 widely participates in and regulates human diseases, including Alzheimer’s disease[6]., breast cancer[7] and atherosclerosis[8].

To date, it has been clarified that inhibiting SHP2 can alleviate the symptoms of psoriasis[9] and Netosis can accelerate the process of psoriasis[10]. However, how Shp2 regulates the process of Netosis in neutrophils is still unknown. Here, we use the imiquimod (IMQ)-induced psoriasis mouse model to investigate the relationship between SHP2 and NETosis in neutrophils. In addition to that, we also used RNA-seq and single-cell RNA-seq technology for better experimental guidance. Based on these data, we propose that SHP2 in neutrophils promotes the release of NETs through the ERK5 pathway which aggravate psoriasis.

## Results

### SHP2 expression by neutrophils is related to develop psoriasis

We collected skin tissue from healthy donor and psoriasis patient, isolated them into single cells and performed single cell RAN-seq to explore the expression of *PTPN11*. We firstly analyzed the expression levels of 107 genes which encoding human PTP phosphatase family members (Supplementary Figure 1), and showed the top ten in Figure 1a. Results showed that the SHP2 coding gene *PTPN11* was highly expressed in skin tissue with psoriasis.

**Figure 1.**
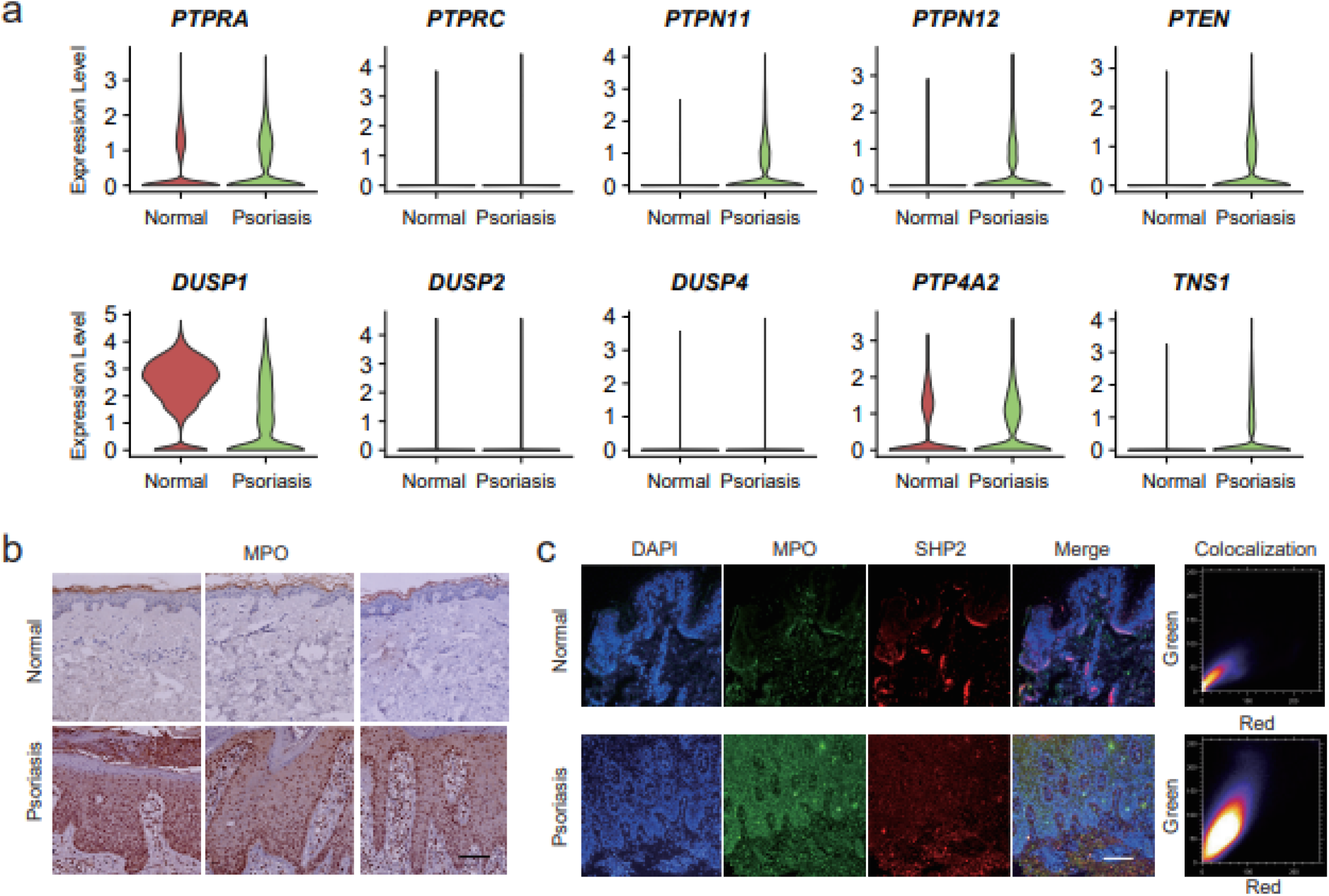
SHP2 expression by neutrophils is indispensable in skin tissue of psoriasis. (a) The violin plot of the top ten expression abundance of 107 PTPs. (b)Neutrophil infiltration was determined by evaluating the intensity of MPO staining of healthy (n=8) and psoriasis patients (n=13) donors, scale bar: 400 μm. (c) Representative immunofluorescence images staining for MPO (green), SHP2 (red) and DAIP (blue) in healthy donors (n=8) and psoriasis patients (n=13) skin section, scale bar: 100 μm.

To determine the function of neutrophils in the pathogenesis of psoriasis, we performed immunohistochemical staining of the skin from patients with psoriasis and healthy donors. Results showed that the infiltration of neutrophils in psoriasis lesions was significantly higher than that in healthy donors (Figure 1c). The results in Figure 1d showed that the expression of SHP2 in neutrophils increased significantly, and had a strong colocalization expression with MPO in the skin lesions of psoriasis patients.

### The deficiency of SHP2 in neutrophils improves psoriasis-like phenotype in mice model

To further study the role of SHP2 in psoriasis, we crossed *Shp2*^flox/flox^ mice with S100a8cre mice and constructed Cre-driven SHP2 deletion in neutrophils. S100a8^cre^-*Shp2*^flox/flox^ (SHP2^N^ KO) mice, and *Shp2*^flox/flox^ (SHP2^N^ WT) mice. The results showed that a series of psoriasis symptoms caused by IMQ on the back of SHP2^N^ WT mice, such as redness and swelling, scaly coverage, and thickening of the epidermis, were significantly improved in SHP2^N^ KO mice (Figure 2a, b). The adapted human clinical Psoriasis Area and Severity Index (PASI) score of the IMQ-treated SHP2^N^ KO mice was also significantly lower than IMQ-treated SHP2^N^ WT mice (Figure 2c). Accordingly, the increase of epidermal thickness caused by IMQ was also alleviated in SHP2^N^ KO mice (Figure 2d). In addition, the expression of inflammatory factors TNF-*α*, IL-1*β*, IL-6, IL-17a, and CXCL15 were significantly reduced in SHP2^N^ KO mice skin (Figure 2e). We injected SHP099 (10 mg/kg) which is an effective SHP2 inhibitor into mice intraperitoneally, and the results were also in line with the previous experimental conclusions: after inhibiting SHP2, the symptoms of psoriasis induced by IMQ were improved (Supplementary Figure 2). In summary, our data indicate that the deletion of SHP2 in neutrophils or inhibition can alleviate IMQ-induced psoriasis symptom.

**Figure 2.**
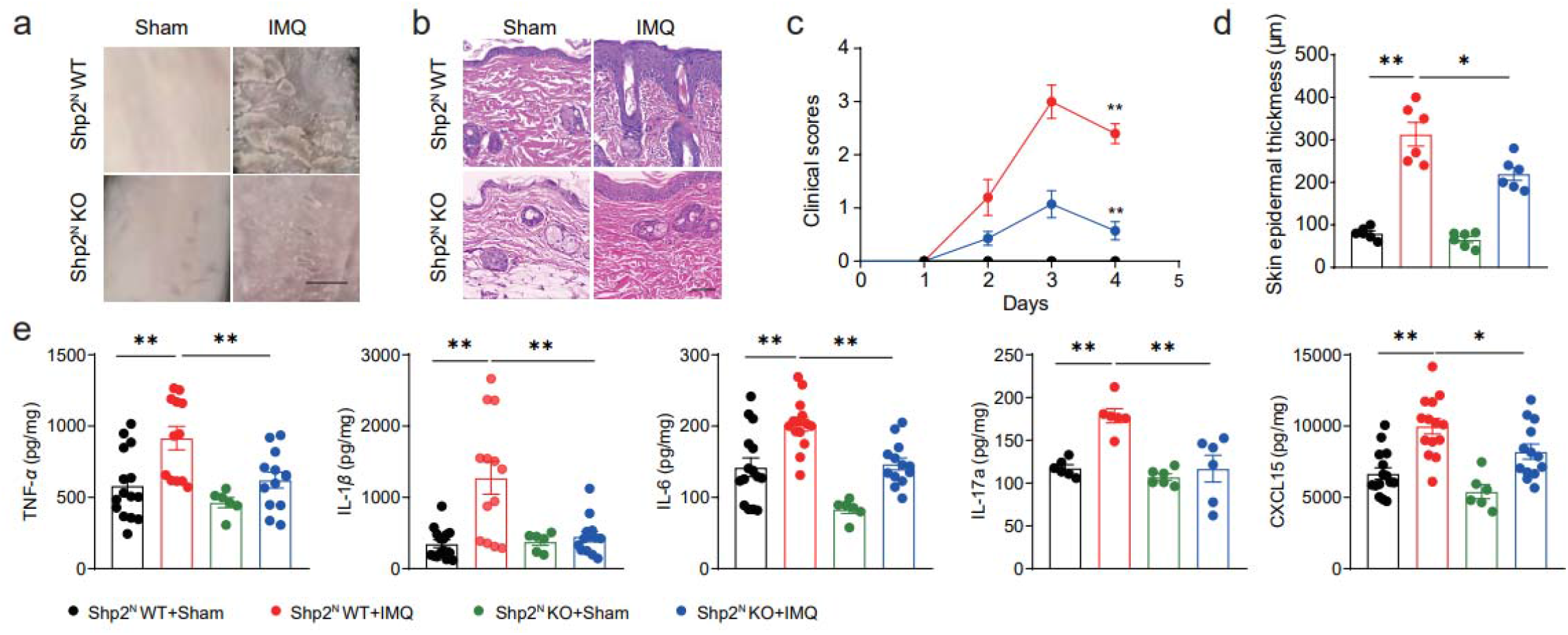
SHP2 deficiency in neutrophil alleviates psoriasis-like phenotype in the IMQ-induced murine model. (a) Phenotypic presentation of back skin sections of sham- or IMQ-treated SHP2N KO and WT mice. (b) The hematoxylin and eosin (H&E) staining of back skin sections of sham- or IMQ-treated SHP2N KO and WT mice, scale bar: 200 μm. Clinical scores (c) or epidermal thickness (d) or of mice dorsal skin. (e) Dorsal skin was infiltered with TNF-α, IL-1β, IL-6, IL-17a and CXCL15 evaluated by ELISA.

### NETs are produced in psoriasis patients and can affect SHP2

To explore the relationship between psoriasis and NETs, we collected the skin tissues of psoriasis patients and healthy donors, and detected CitH3 (Figure 3a) and PAD4 (Figure 3b) protein expression, which are 2 key proteins formed in the process of NETosis by immunohistochemistry. The results showed that the expression of CitH3 and PAD4 in skin lesions of psoriasis patients increased significantly compared with the skin of healthy donors. In addition, we used immunofluorescence method to obtain images of NE and MPO in skin sections to observe NETs. The results showed that more NETs were produced in the skin of patients with psoriasis (Figure 3c). Next, we quantified dsDNA which can reflect the generation of NETs from the serum of normal people and psoriasis. Results showed that the amount of dsDNA from psoriasis serum samples are sufficiently larger than that from normal people (Figure 3d). Finally, we analyzed genes’ mRNA expression which relates to NETs formation and *PTPN1*1 in human skin tissue. Data from single cell RNA-seq showed that mRNA expression of *ELANE* and *PADI4* increased with the increase of *PTPN11* expression.

**Figure 3.**
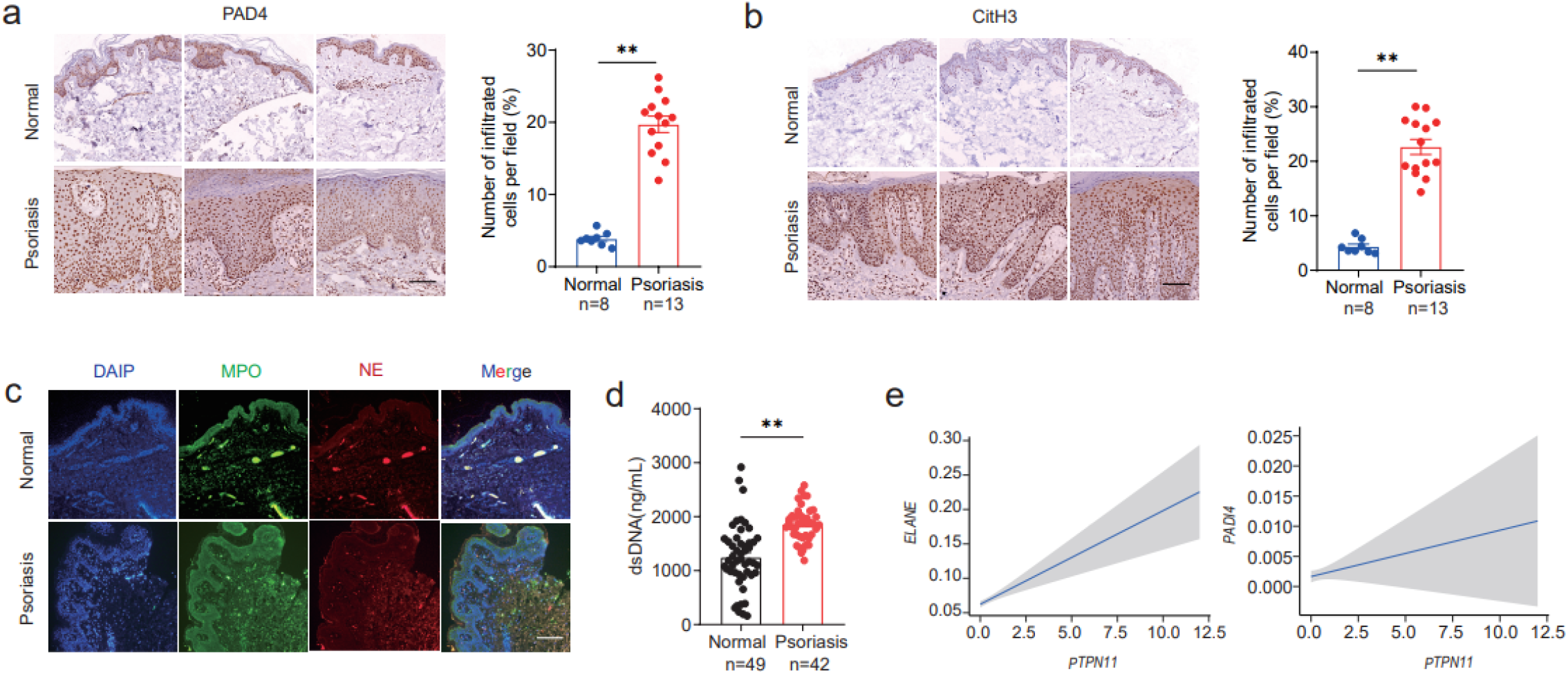
NETs are produced in psoriasis patients. Immunohistochemical staining of skin sections from psoriasis patients and healthy donors with anti-PAD4 (a) or anti-CitH3 (b), scale bar: 400 μm. (c) Immunofluorescence staining of skin sections from psoriasis patients and healthy donors with anti-MPO, anti-NE, and DAPI, scale bar: 400 μm. (d) Quantification of dsDNA in serum samples from healthy and psoriasis donors. (e) Expression level of *PTPN11, ELANE* and *PADI4* in human skin.

These data indicate that pathways involved in the formation of NETs are activated in skin tissues of psoriasis patients, and NETs are generated more if there are more SHP2 expression.

### NETs can mediate the pathogenesis of psoriasis like symptom in mice

PAD4 protein has been reported to be closely related to the formation of NETs, and it was coding by *Padi4* in mice. Next, we constructed *Padi4* knockout (Pad4 KO) mice which were unable to form NETs. To confirm the efficiency of PAD4 deletion, we dissociated the skin of PAD4 KO mice and examined the expression of PAD4 using immunohistochemistry (Figure 4a).

**Figure 4.**
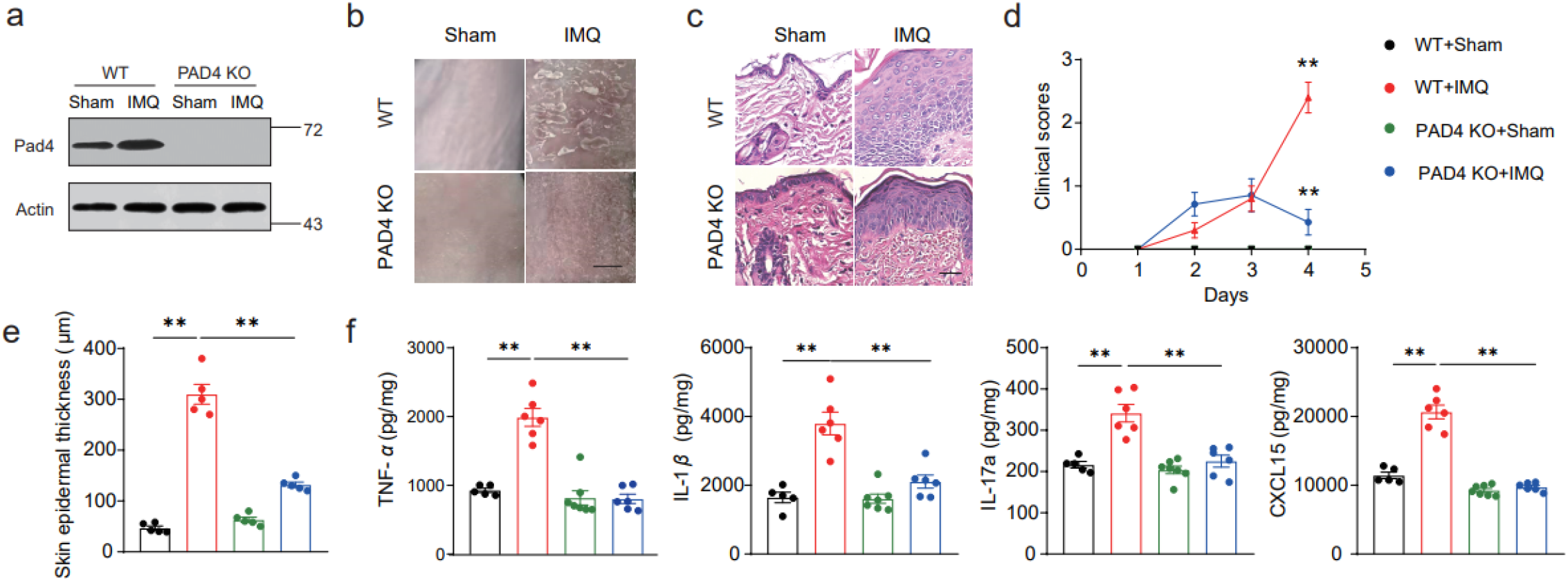
PAD4 deficiency alleviates psoriasis-like phenotype in the IMQ-induced murine model, and led the growth of SHP2. (a) The protein level of PAD4 in mice dorsal skin, tested by western blot. (b) Phenotypic presentation of sham- or IMQ-treated PAD4 KO and WT mice, scale bar: 50mm. (c) H&E staining of back skin sections of sham- or IMQ-treated PAD4 KO and WT mice, scale bar: 200 μm. Clinical scores(d) or epidermal thickness(e) of mice dorsal skin. (f) Dorsal skin was infiltered with TNF-α, IL-1β, IL-17a and CXCL15 evaluated by ELISA.

In the following experiment, PAD4 KO or WT mice were randomly divided into 2 groups, which treated with equal amounts of imiquimod or control cream for 3 days, and skin tissue was obtained on day 4. Compared with WT mice, the deletion of *Padi4* significantly reversed the psoriasis-like symptoms caused by imiquimod. Scaling was significantly improved in PAD4 KO mice (Figure 4b). In addition, the epidermal hyperplasia was also significantly reduced proven by epidermal thickness and hematoxylin-eosin staining as we can see in Figure 4c, e. What’s more, the PASI score of IMQ treated WT mice continued to increase and peaked on day 4, while the PASI score of PAD4 KO mice continually kept on a low value, and decreased on day 4 (Figure 4d). Furthermore, we tested the expression of psoriasis-related inflammatory cytokines in mice skin. The results showed that *Padi4* gene knockout effectively reversed the overexpression of inflammatory factors TNF-α, IL-1β, CXCL15 and IL-17a on the skin of mice caused by IMQ (Figure 4f). These data suggest that reduced level of NETs has an effective fact on ameliorating IMQ-induced psoriatic skin disorders. In conclusion, decreasing NETs can alleviate the development of IMQ-induced psoriasis mice model.

### Single-cell RNA-seq reveals the inhibition of SHP2 can decrease neutrophils

We collected the skin tissues of mice which treated with IMQ or SHP099 or none, dissociated them into single cells and performed the single-cell RNA-seq (Figure 6a). Unbiased clustering revealed 5 cell clusters (Figure 6b), and we can see the cell distribution as shown in Figure 6c. According to specific marker genes, we defined these clustered cell populations as fibroblasts, immune cells, keratinocytes, Schwann cells and smooth muscle cells (Figure. 6d). Among them, immune cell subsets were defined as macrophages, T cells, neutrophils and natural killer T cells (Figure 6e). We found that the proportion of neutrophils in immune cells increased significantly after IMQ treatment, and decreased after SHP099 treatment (Figure 6f). Next, we analyzed neutrophil related genes. Results showed that neutrophil infiltration occurred in the skin tissues after IMQ treatment, and SHP099 significantly reversed this phenomenon (Figure 6g, h).

**Figure 5.**
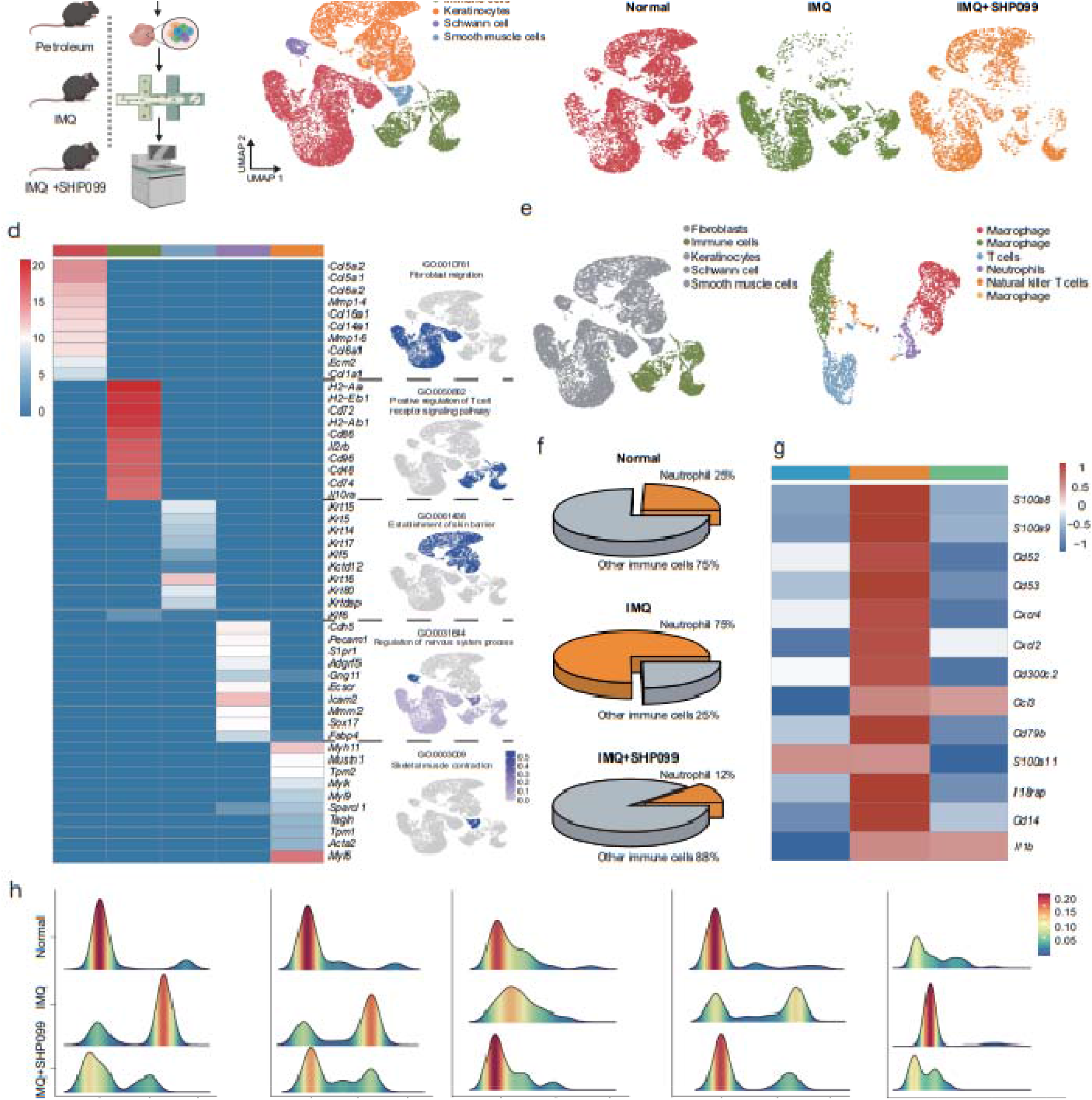
Single-cell RNA-seq reveals the inhibition of SHP2 can decrease neutrophils. (a) Schematic representation of the experimental procedure. (b) Unbiased clustering reveals 5 cellular clusters. (c) Cell atlas of mice skin tissue. (d) Heat map image of functional gene set and cell atlas of each cell population. (e) Unbiased clustering of immune cells. (f) Pie chart of the proportion of neutrophils to immune cells. (g) Heat map image of neutrophil related functional gene sets; (h) Summit plot of neutrophil related functional genes.

**Figure 6.**
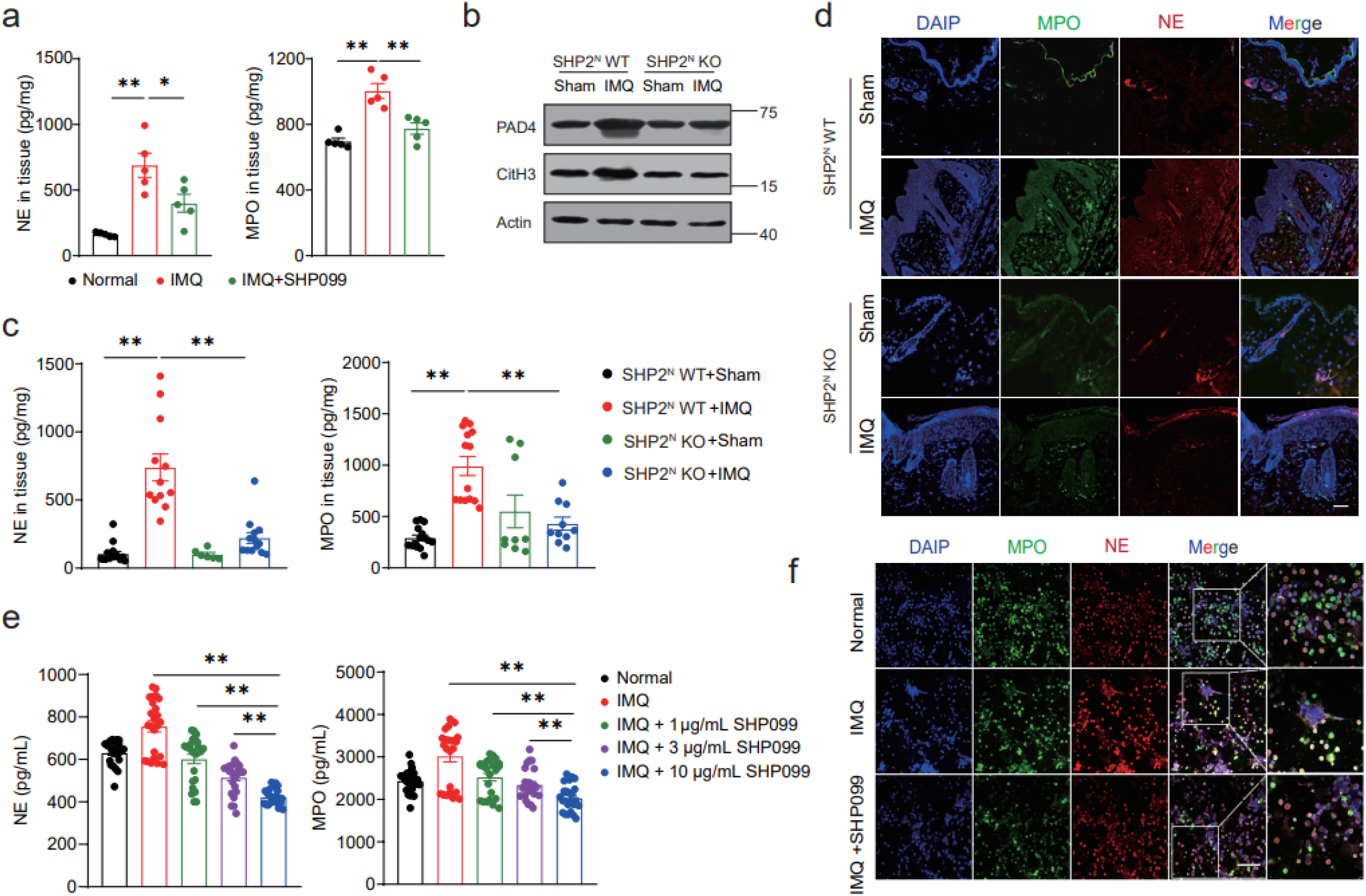
Inhibition of SHP2 expression in neutrophils can inhibit the formation of NETs. (a) The protein expression of NE and MPO in the IMQ or SHP099 treated mice. (b) The protein level of CitH3 and PAD4 of mice dorsal skin, tested by western blot. (c) The protein expression of NE and MPO of mice dorsal skin tested by ELISA. (d) Representative immunofluorescence images staining for MPO (green), NE (red) and DAIP (blue) of mice skin section, scale bar: 200 μm. (e) The protein expression of NE and MPO in neutrophils extracted from peripheral blood. (f) Representative immunofluorescence images staining for MPO (green), NE (red) and DAIP (blue) of neutrophils extracted from peripheral blood, scale bar: 100 μm.

### Inhibition or deletion of SHP2 can inhibit the formation of NETs

In order to evaluate the effect of SHP2 in NETs formation, we firstly explored NE and MPO expression in IMQ or SHP099 treated mice. Results showed that the expression of protein NE and MPO were decreased by SHP2 inhibition (Figure 6a). Then, we used neutrophils specific SHP2 knockout mice to make some future exploration. ELISA and immunofluorescence were used to test the expression level of NE and MPO of mice dorsal skin. The results showed that IMQ significantly increased the expression of NE and MPO, and *SHP2* specific knockout in neutrophils had revered this situation. The same trend was proved in the expression of CitH3 and PAD4 proteins detected by Western blot (Figure 6b, c, d). Next, we extracted neutrophils from peripheral blood of healthy donors, and then processed them with IMQ and/or SHP099. The results showed that IMQ treatment significantly increased the expression of NE and MPO, while SHP099 effectively inhibited the expression of these two proteins in a dose-dependent manner (Figure 6e, f). These results preliminarily confirmed that NETs which are involved in the progression of psoriasis could be down regulated by SHP2 inhibition or deletion.

### SHP2 promotes the formation of NETs through the ERK5 pathway

To verify the regulatory relationship between SHP2 and NETs, we isolated neutrophils from human peripheral blood and treated the cells with IMQ or/and SHP099. RNA sequencing showed that: genes encoding proteins involved in the formation of NETs were highly expressed after IMQ treatment, such as *MPO, Elane*, and *PADI4*; Genes encoding inflammatory factor, such as *IL1B* and *IL6* were highly expressed too after IMQ treatment, whereas SHP099 reversed the phenomenon. In addition to that, genes which relate to the formation of NETs were enriched[11], such as MAPK family, *AKT1, AKT2, GSDMD* and so on (Figure 7a). Previous studies have reported that the MAPK pathway is involved in the formation of NETs. Therefore, we used qPCR to explore four major MAPK family classifications: ERK, ERK5, JNK, p38 MAPK, which was encoded by *MAPK1, MAPK7, MAPK8, MAPK14* respectively. We found that *MAPK7* was significantly overexpressed after IMQ treatment (Figure 7b). Then, we treated neutrophils with IMQ or/and SHP099 for 30 min 60 min or 120 min. We found that SHP099 decreased the high expression of *MAPK7* caused by IMQ, and the decrease was more significant with longer treating time (Figure 7c).

**Figure 7.**
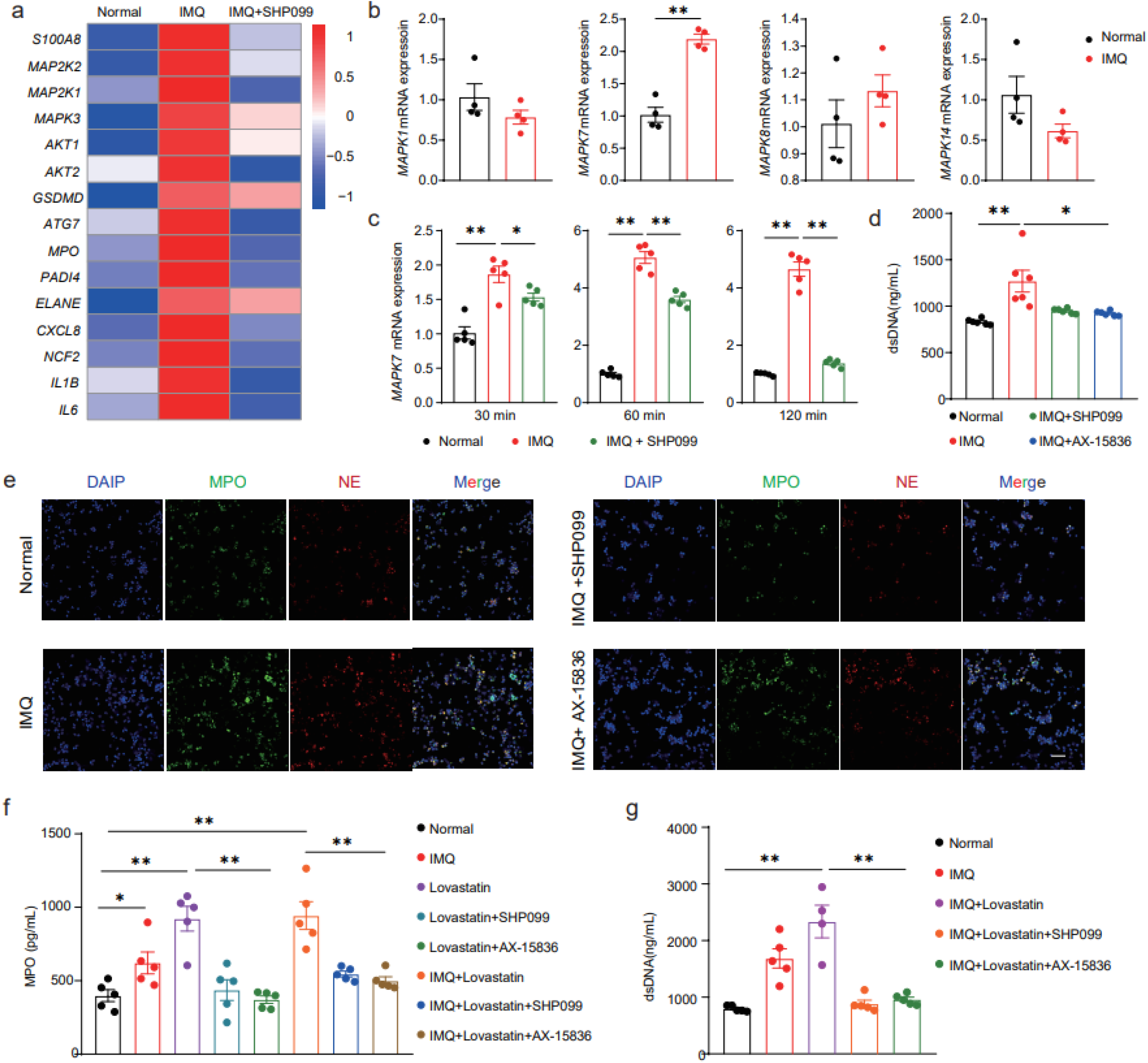
SHP2 promotes the formation of NETs through the ERK5 pathway. (a) Heat map image of microarray analysis of RNA from human neutrophils (n=3/group). (b) Expression levels of representative MAPK family genes in neutrophils. (c) Expression levels of *MAPK7* in neutrophils in different time limit. (d) Quantification of dsDNA in neutrophils. (e) Representative immunofluorescence images staining for MPO (green), NE (red) and DAIP (blue) of neutrophils, scale bar: 30 μm. (f) The protein expression of MPO in neutrophils tested by ELISA. (g) Quantification of dsDNA in neutrophils.

Next, we treated neutrophils with ERK5 inhibitor: AX-15836 and found that, the formation of NETs was inhibited which similar with the effect of SHP099 (Figure 7d, e). Finally, we treated neutrophils with lovastatin, which has been reported to elevate SHP2 activity in cells[12], and showed that lovastatin was able to induce a high expression of NETs, whereas after inhibition of SHP2 or ERK5, NETs were no longer produced in large amounts. This conclusion was supported by the MPO protein expression measured by ELISA, as well as dsDNA quantitative detection (Figure 7f, g). In conclusion, SHP2 may promote the formation of NETs through the ERK5 pathway.

## Discussion

Accumulating evidence indicates that SHP2 is associated with autoimmune diseases, such as in systemic lupus erythematosus and rheumatoid arthritis. In these situations, SHP2 usually plays a role in aggravating disease. Psoriasis, as a kind of autoimmune disease, has caused serious harm to the body and psychology of patients[13]. The relationship between SHP2 and psoriasis has also been elucidated in 2020, in which, SHP2 promoted the transport of TLR7 from the Golgi to the endosome, and led the interaction with and dephosphorylation of TLR7 at Tyr1024, thereby promoted TLR7 ubiquitination and psoriatic skin inflammation [6]. In addition, the research of NETs in the field of psoriasis is also quite popular. NETs formation is increased in both peripheral blood and lesion skin of psoriasis patients and correlates with disease severity[14].However, whether there is a regulatory relationship between SHP2 and NETs in neutrophils, and how SHP2 could influence NETs to promote psoriasis await future investigation. Our results suggest that, in neutrophils, SHP2 acts as a key factor in the pathogenesis of psoriasis and can promote the generation of NETs and increase the expression of inflammatory cytokines associated with psoriasis through the ERK5 pathway. This study suggests that the ERK5 pathway is a potential therapeutic target for the treatment of psoriasis.

The association of SHP2 with various skin diseases has been extensively studied. First, for the effects of SHP2 with skin cancer. SHP2 was primarily responsible for UVB mediated dephosphorylation of STAT3 and may act as a protective mechanism against UV skin carcinogenesis[15]. Zhang et al proposed SHP2 promotes melanoma cell viability, migration, and colony formation by activating ERK1/2 and Akt signaling pathway, so SHP2 inhibitors could act as a potential novel therapeutic agent for melanoma treatment[16]. What’s more, SHP2 is involved in the process of skin fibrosis. Distler et al characterized SHP2 as a TGFβ’s molecular checkpoint, which could block JAK2 / STAT3 signaling. SHP2 was able to serve as a potential target for the treatment of skin fibrosis[17].

Among the diseases listed above, suppression of SHP2 expression is unspecific. The investigation of SHP2 expression specifically in certain kinds of cells has great research potential. SHP2 specifically expressed in macrophage, and could positively regulate oxidative reaction through its’ phosphatase activity[18]. What’s more, Shp2 could also regulates the responsiveness of macrophage to interleukin-10 which influenced Intestinal health[19]. It is well known that neutrophils play an important role in body immunity. They are the most common leukocytes in the blood and one of the earliest responsible immune cells to fight infection. In a mouse model of LPS induced ALI, neutrophils can play a key role in the pathogenesis of ALI/ARDS by promoting inflammation and injury to the alveolar microenvironment. Research found out that targeted the inhibition of SHP2 in neutrophils could alleviate lung inflammation[20].

In 2018, Muraro et al. revealed the relationship between MAPK family and NETosis: respiratory syncytial virus is the main cause of respiratory diseases in infants. It induces netosis through PI3K/Akt, ERK and p38 MAPK signal transduction[21]. Here We verified that SHP2 could promote NETosis in neutrophils through the ERK5 pathway which leads to psoriasis. Our study provides evidence for the role of SHP2 in NETosis in the progression of psoriasis, and SHP2 may be a potential therapeutic target for the treatment of psoriasis.

## Materials and methods

### Animal experiments

All the experimental animals were purchased from GemPharmatech Co. Ltd., and animal were maintained in specific pathogen-free conditions at GemPharmatech Co. Ltd and Experimental Animal Center at Nanjing University. *Shp2* flox/flox mice were crossed with S100a8-cre mice on the C57BL/6 background for more than two generations to generate Shp2 flox/flox S100a8-cre mice which SHP2 was conditional knocked out in neutrophils. we also constructed *Padi4* knockout on the C57BL/6 background.

Mice were shaved and treated with imiquimod (H20160079, 3M Pharmaceuticals, USA) or petroleum successively. For different experimental needs, 10 mg/kg SHP099, or physiological saline solution was injected intraperitoneally (i.p.). Serum was isolated from the blood of mice which anesthetized with 0.05 mg/kg pelltobarbitalum natricum, and followed by retro-orbital sinus taken. Skin and serum were stored at - 80 □ for future use. All the procedures were carried out in accordance with the Guide for the Care and Use of Laboratory Animals (National Institutes of Health, USA) and ethical regulations of Nanjing university to guarantee animal welfare.

### Human specimens

Psoriasis patients and normal healthy donors’ skin tissue were taken to make paraffin sections. Sample acquisitions were approved by the Ethics Committee of West China Hospital. In process, we obeyed the declaration of Helsinki Principles.

### PASI score

The changes of mice dorsal skin were observed and calculated according to the PASI scoring standard for the degree of erythema, scaling and infiltration. Then the points of the three items were summed to obtain a total score. The scoring standard refers to the “Severity Severity Scoring Table of Mouse Psoriasis Model” to compare the PAIS scores of the three groups of mice. Each index ranges from (0 to 4) points, 0 points: none; 1 point: mild; 2 Points: moderate; 3 points: obvious; 4 points: extremely obvious.

### Neutrophils isolation

Ficoll-Hypaque density gradient centrifugation were used to isolate neutrophils from the peripheral blood of healthy donors. According to the instructions of Human Neutrophil Isolation Kit (LZS11131, Haoyang Biology, China), neutrophil separation solution of different concentration gradients has been already added in advance to the sterile silicified centrifuge tube. Peripheral blood and hydroxyethyl starch were mixed and then gently superimposed on the separation solution. After 20 min’s centrifugation at 800 g, neutrophils were pipetted out, followed by washing, centrifugation, and resuspend. Neutrophils were planted in 9 cm cell culture dishes with RPMI 1640 Medium (01-100-1A, Biological Industries, Israe), 10 % fetal bovine serum (2033119, Biological Industries, Israe) and 1% penicillin-streptomycin (CO222, Beyotime, China). Cells grew in an incubator at 37 □ with 5% CO_2_.

### Immunohistochemistry

The immunohistochemistry experiment was performed according to the instructions of immunohistochemistry detection kit (KIHC-5, Proteintech Group, USA). Skins were embedded in paraffin and made into 5 μm slides. Slides were treated with 5% sodium citrate buffer (P0081, Beyotime, China) and 2% hydrogen peroxide for antigen retrieval and endogenous peroxidase activity blocking. 3% goat serum was used to block nonspecific binding sites of antibodies. Then the slides were incubated with the primary antibody: rabbit anti-myeloperoxidase antibody (ab208670, Abcam, USA), mouse anti-neutrophil elastase antibody (sc-55549, Santa Cruz Biotechnology, USA) at 4 °C in a humidified chamber overnight. Next day, slides were treated with anti-mouse/rabbit immunohistochemistry detection kit (PK10006, Proteintech, USA). The hematoxylin was used to observe the nucleus. Finally, the slides were imaged by a light microscope (IX61, Olympus, Japan).

### Immunofluorescence

Immunofluorescence staining was performed on tissue and cell samples. For tissue samples, slides were deparaffinized, rehydrated, and treated with sodium citrate buffer for antigen retrieval. Slides were incubated with 3% goat serum for 30 min followed by primary antibodies. Cell samples were fixed in poly-L-lysine polylysine (P2100, Solarbio, China), followed by permeabilization and nonspecific binding sites blocking. Samples were treated at 4 °C overnight with the following primary antibodies: rabbit anti-myeloperoxidase antibody (ab208670, Abcam, USA), mouse anti-neutrophil elastase antibody (sc-55549, Santa Cruz Biotechnology, USA), goat anti-shp2 antibady (ab9214, Abcam, USA), rabbit anti-pad4 antibody (ab214810, Abcam, USA), rabbit anti-histone H3 (citrulline R2) antibody (ab174992, Abcam, USA). Next day, Samples were incubated with an Alexa fluorescein-labeled secondary antibody for 2 h. DAPI (C1002, Beyotime, China) was used to stain cell nuclei. Then samples were imaged by an inverted confocal microscope (LSM880 with airyscan, Carl Zeiss, Germany).

### Histopathologic assessment

Slides were deparaffinized, rehydrated, and treated with hematoxylin solution and eosin staining according to the instruction of Hematoxylin-eosin staining kit (G1005, Servicebio, China) to evaluate pathological changes in skin tissue. Then slides were dehydrated and sealed with neutral balsam.

### Western blotting assay

Protein samples were prepared by tissue lysate and BCA protein assay. 15 μL protein of each sample was loaded and separated by 10% sodium dodecyl sulfate-polyacrylamide gel electrophoresis (SDS-PAGE) and transferred onto polyvinylidene fluoride (PVDF) membranes (IPVH00010, Merck Millipore, Germany). The membranes were washed with 5% nonfat milk for 2 h to block the non-specific binding sites and then treated with primary antibody: rabbit anti-pad4 antibody (ab214810, Abcam, USA), rabbit anti-histone H3 (citrulline R2) antibody (ab174992, Abcam, USA), mouse anti–β-actin antibody (M20011, abmart, China) overnight at 4 °C. The secondary antibody was incubated with the membranes for 2 hours. Signals were detected and exposed by using LumiGLO^®^ Reagent (#7003, Cell Signaling Technology, USA).

### dsDNA quantification

Quant-it PicoGreen dsDNA Assay kit (P7581, Thermo Fisher Scientific, USA) was used to quantify cfDNA in mice serum. Samples, TE buffer, and Quant-iT PicoGreen reagent were added into a 96-Well Black Opaque Plate. Fluorescence signals were detected by a microplate fluorescence reader (Safire, Tecan, Switzerland) with an excitation wavelength of 480□nm and an emission wavelength of 520□nm.

### ELISA

The expression of IL-1β, TNF-α, IL-6, CXCL-15, NE, and MPO was detected by DuoSet ELISA kit (R&D System, USA). Briefly, the capture antibodies were coated in a 96-well enzyme linked immunoassay plate (Costar 3590, Corning Incorporated, USA) at first day in room temperature. After treated with washing buffer, skin lysate and serum were incubated in the plate for 2 h, then the detection antibodies were treated, absorbance at 450 nm was detected by a microplate (Safire, Tecan, Switzerland).

IL-17A.

### Quantitative PCR

Trizol was used to extract total RNA (108-95-2, Takara, China). After quality control, we quantified 1 μg RNA, and reverse transcribed it to synthesize single stranded cDNA. We performed quantitative PCR for CFX 100 (bio rad, Hercules, CA) cyclers using the primers listed in Supplementary Table 1. The procedure of the amplification program was: 95 °C for 2.5 min, 95 °C for 15 s, 60 °C for 30 s, for a total of 44 cycles. Dissociation curves were analyzed at the end of amplification. RNA expression levels of GAPDH were used for normalization.

### Statistical analysis

All data are expressed as the mean ± SEM and were analyzed by GraphPad Prism 8. 0. Student’s *t* test and one-way ANOVA test were used for statistical analyses of the data. Differences were considered significant at \**P* < 0. 05 or \*\**P* < 0.01.

## Supporting information

Supplementary figures and will be used for the link to the file on the preprint site

## Data Availability

All data produced in the present study are available upon reasonable request to the authors

## Acknowledgments

We thank Prof. Bisen Ding (Sichuang University) for providing S100a8-Cre mice. This work was supported by National Natural Science Foundation of China (Nos. 81872877, 81730100, 91853109, 81673436, 81803142), and Mountain-Climbing Talents Project of Nanjing University.

## Figures and Tables

**Supplementary Figure 1.**
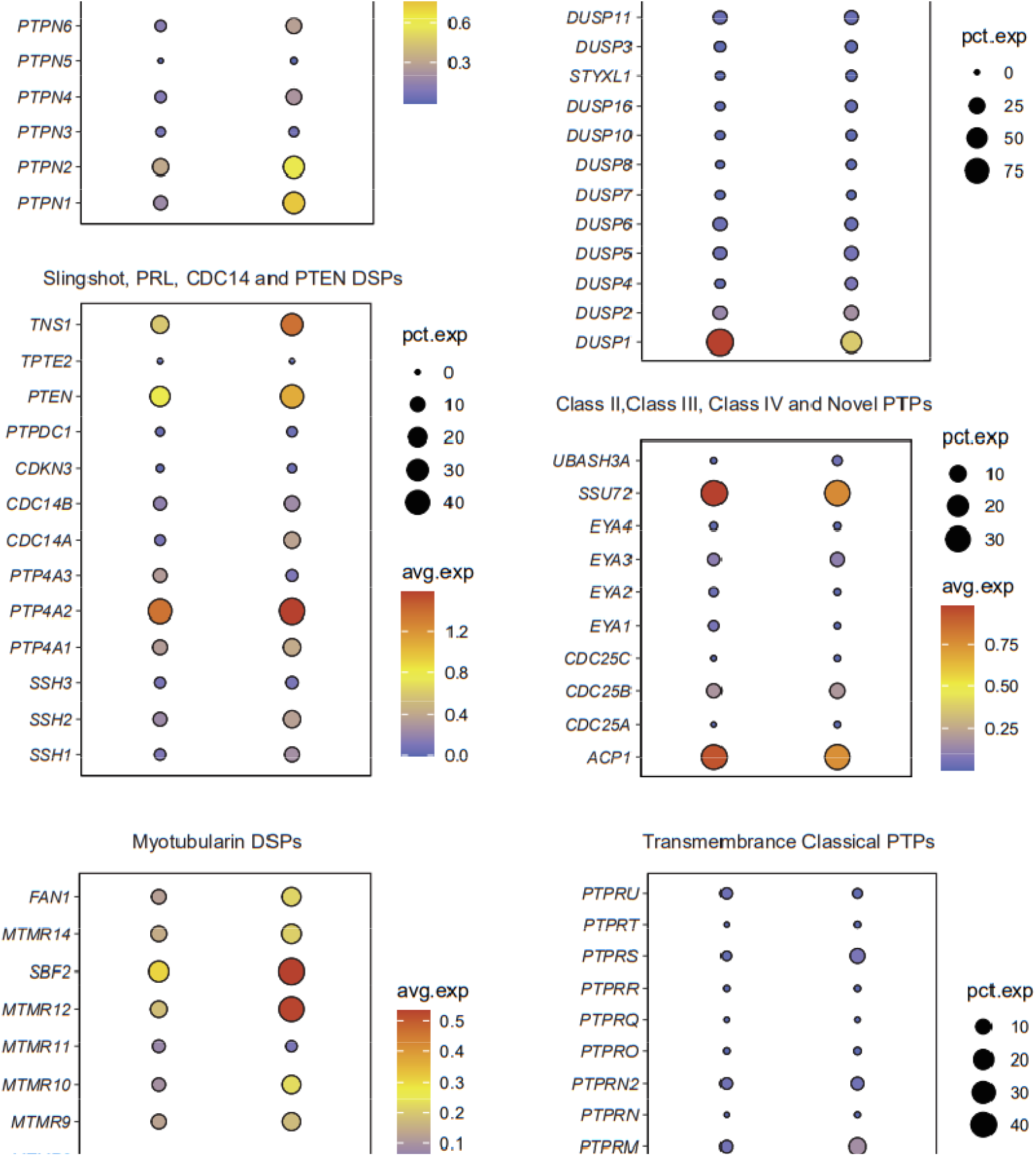
Bubble chart of the expression abundance of 107 PTPs which are classified as 6 subtypes.

**Supplementary Figure 2.**
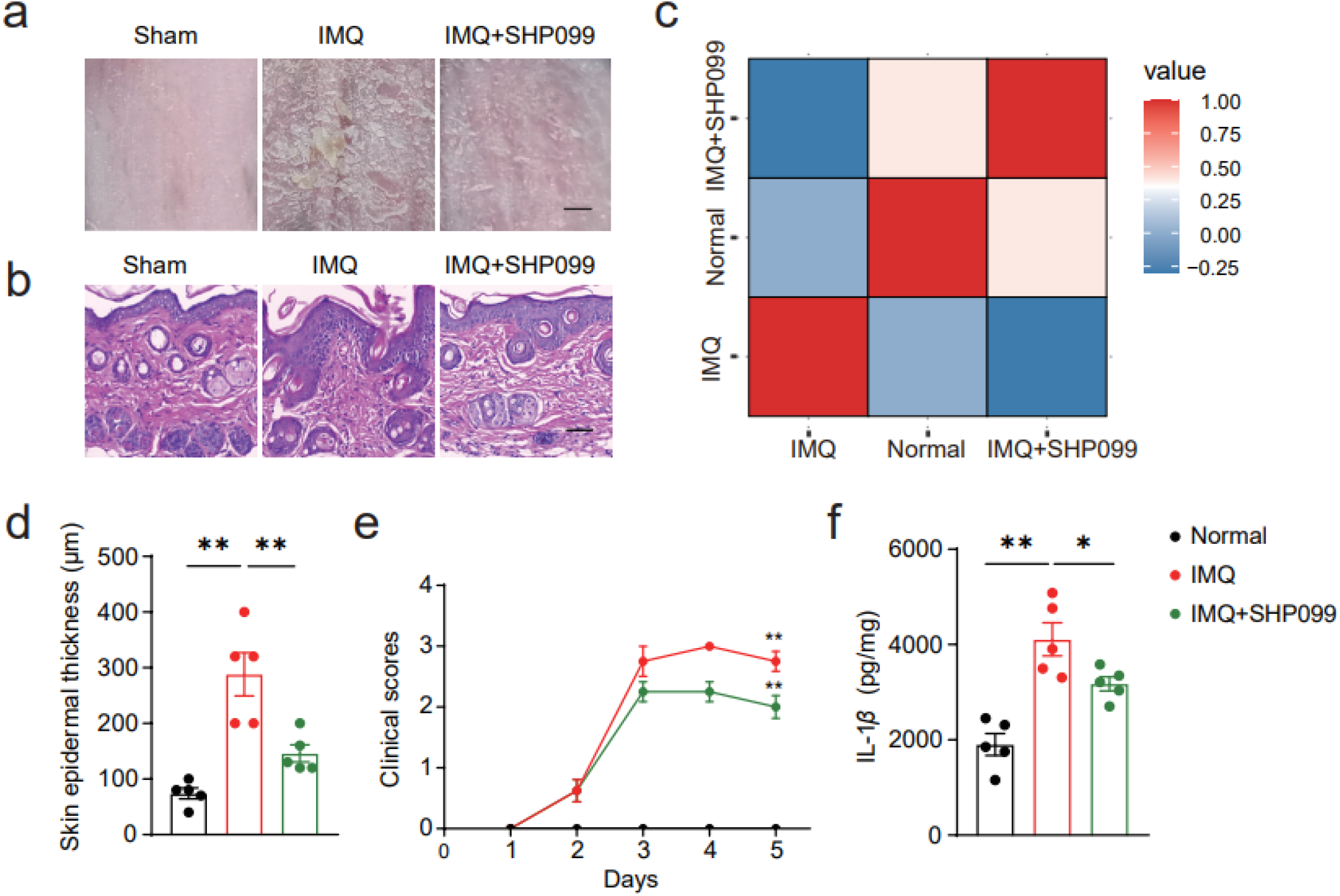
Inhibition of SHP2 can alleviate psoriasis like symptoms in mice. (a) Phenotypic presentation of back skin sections of sham-, IMQ- or/and SHP099-treated mice. (b) The hematoxylin and eosin (H&E) staining of back skin sections of mice, scale bar: 200 μm. (c) Pearson correlation between samples. Epidermal thickness (d) or clinical scores (e) of mice dorsal skin. (e) Dorsal skin was infiltered with IL-1β evaluated by ELISA.

**Supplementary Figure 3.**
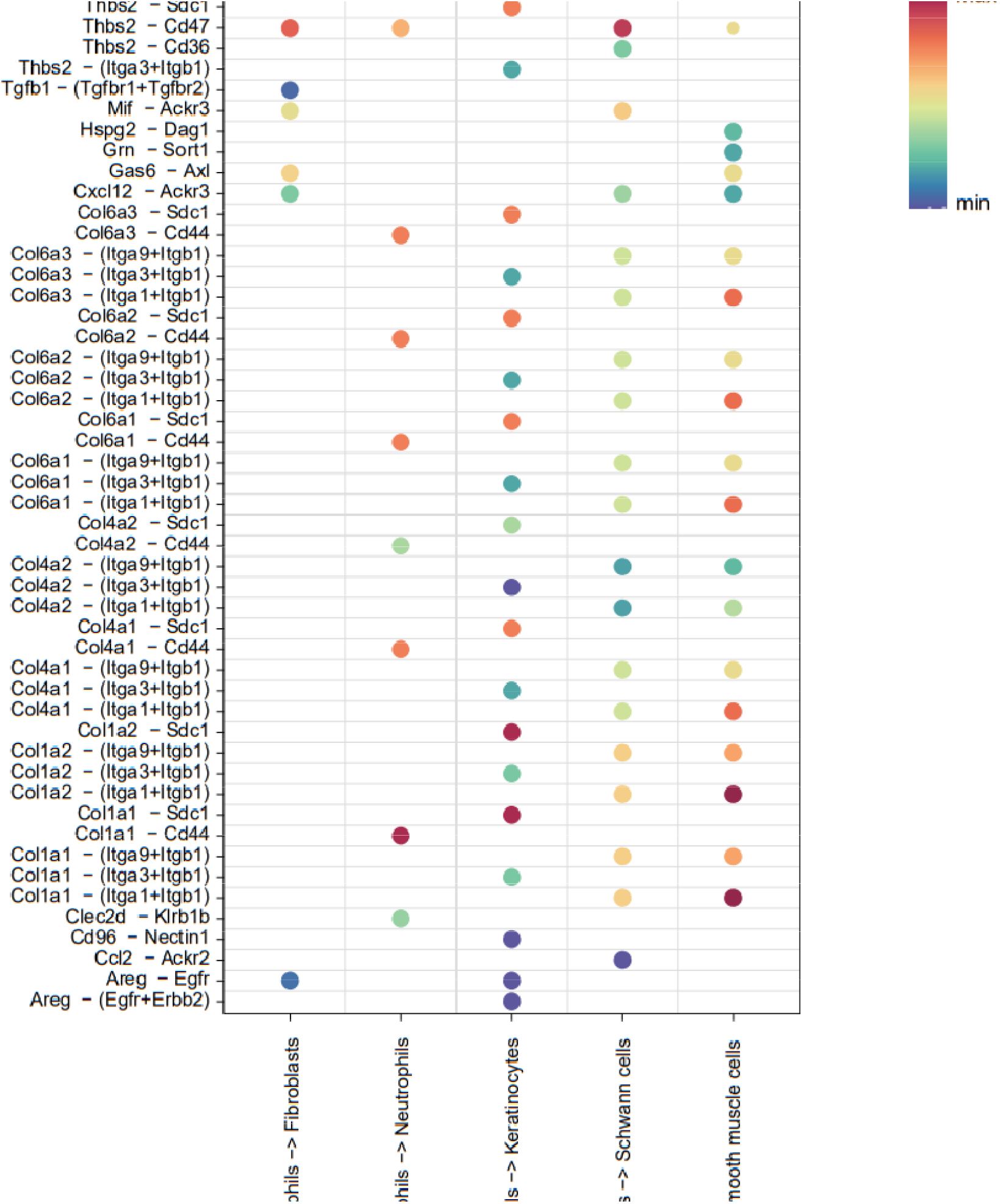
Cell interaction plot of mice skin tissue.

**Supplementary Table 1.**
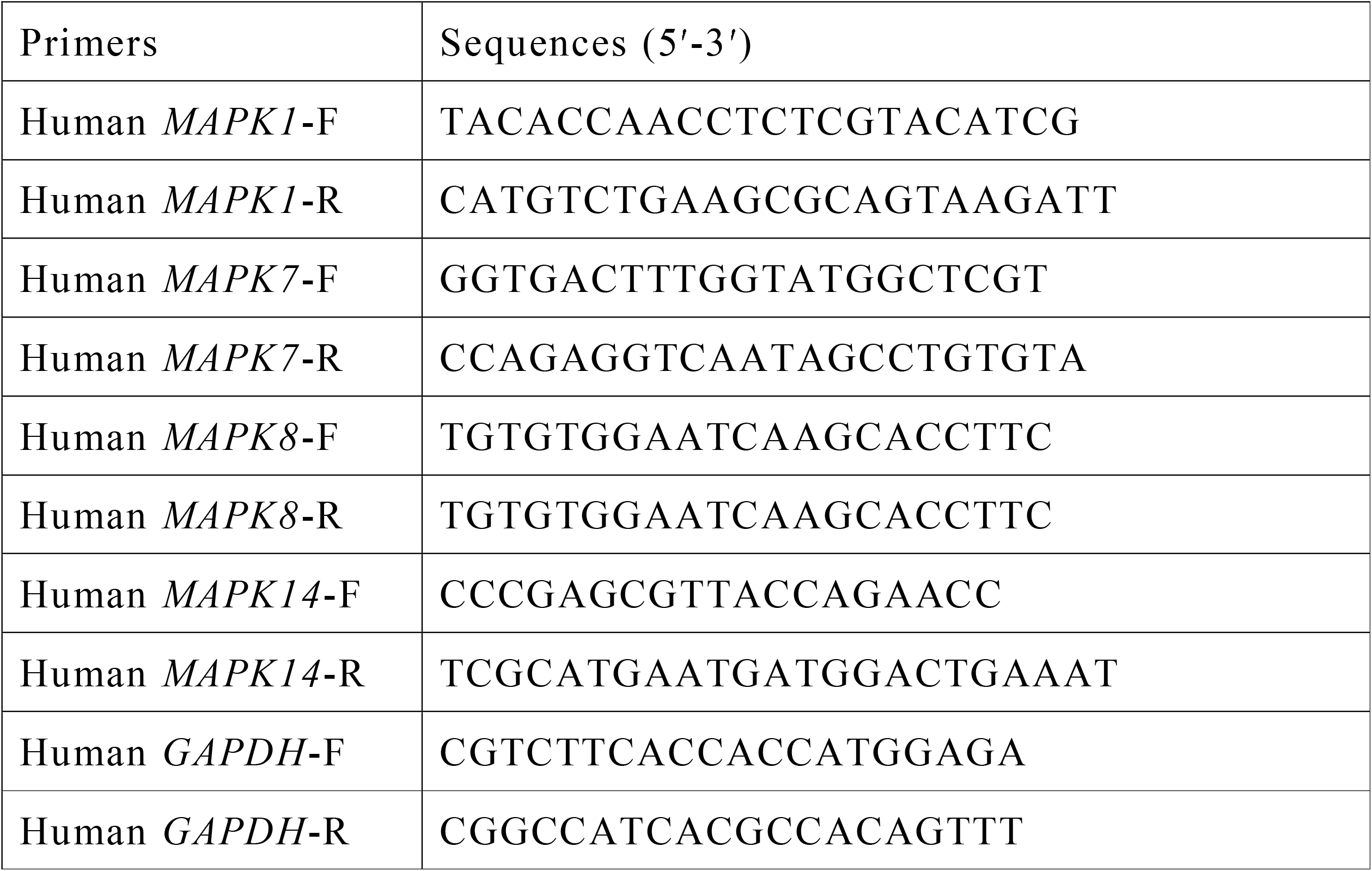
Primers for quantitative PCR analysis.

## References

[1] An emerging role for neutrophil extracellular traps in noninfectious disease”. Nature Medicine. 23 (3): 279–287. doi:10.1038/nm.4294. ISSN 1078-8956. PMID 28267716.

[2] Thiam HR, Wong SL, Qiu R, Kittisopikul M, Vahabikashi A, Goldman AE, Goldman RD, Wagner DD, Waterman CM. NETosis proceeds by cytoskeleton and endomembrane disassembly and PAD4-mediated chromatin decondensation and nuclear envelope rupture. Proc Natl Acad Sci U S A. 2020 Mar 31;117(13):7326–7337. doi: 10.1073/pnas.1909546117. Epub 2020 Mar 13. PMID: 32170015; PMCID: PMC7132277. rupture.” Proceedings of the National Academy of Sciences 117.13 (2020): 7326-7337.

[3] Hou J, Xia Y, Jiang R, Chen D, Xu J, Deng L, Huang X, Wang X, Sun B. PTPRO plays a dual role in hepatic ischemia reperfusion injury through feedback activation of NF-κB. J Hepatol. 2014 Feb;60(2):306–12. doi: 10.1016/j.jhep.2013.09.028. Epub 2013 Oct 12. PMID: 24128416.

[4] Xu Y, Zhang Y, Yang Y, Liu Y, Tian Q, Liu P, Ding Z, Cheng H, Zhang X, Ke Y. Tyrosine phosphatase Shp2 regulates p115RhoGEF/Rho-dependent dendritic cell migration. Cell Mol Immunol. 2021 Mar;18(3):755–757. doi: 10.1038/s41423-020-0414-y. Epub 2020 Apr 29. PMID: 32350404; PMCID: PMC8027028.

[5] Liu Q, Qu J, Zhao M, Xu Q, Sun Y. Targeting SHP2 as a promising strategy for cancer immunotherapy. Pharmacol Res. 2020 Feb;152:104595. doi: 10.1016/j.phrs.2019.104595. Epub 2019 Dec 12. PMID: 31838080.

[6] Su C, Sun F, Cunningham RL, Rybalchenko N, Singh M. ERK5/KLF4 signaling as a common mediator of the neuroprotective effects of both nerve growth factor and hydrogen peroxide preconditioning. Age (Dordr). 2014;36(4):9685. doi: 10.1007/s11357-014-9685-5. Epub 2014 Jul 12. PMID: 25015774; PMCID: PMC4150906.

[7] Montero JC, Ocaña A, Abad M, Ortiz-Ruiz MJ, Pandiella A, Esparís-Ogando A. Expression of Erk5 in early stage breast cancer and association with disease free survival identifies this kinase as a potential therapeutic target. PLoS One. 2009;4(5):e5565. doi: 10.1371/journal.pone.0005565. Epub 2009 May 15. PMID: 19440538; PMCID: PMC2678256.

[8] Heo KS, Cushman HJ, Akaike M, Woo CH, Wang X, Qiu X, Fujiwara K, Abe J. ERK5 activation in macrophages promotes efferocytosis and inhibits atherosclerosis. Circulation. 2014 Jul 8;130(2):180–91. doi: 10.1161/CIRCULATIONAHA.113.005991. Epub 2014 Apr 28. PMID: 25001623; PMCID: PMC4439099.

[9] Zhu, Yuyu, et al. “Inhibition of SHP2 ameliorates psoriasis by decreasing TLR7 endosome localization.” medRxiv (2020).

[10] Di Domizio J, Gilliet M. Psoriasis Caught in the NET. J Invest Dermatol. 2019 Jul;139(7):1426–1429. doi: 10.1016/j.jid.2019.04.020. PMID: 31230639.

[11] Muraro SP, De Souza GF, Gallo SW, Da Silva BK, De Oliveira SD, Vinolo MAR, Saraiva EM, Porto BN. Respiratory Syncytial Virus induces the classical ROS-dependent NETosis through PAD-4 and necroptosis pathways activation. Sci Rep. 2018 Sep 21;8(1):14166. doi: 10.1038/s41598-018-32576-y. PMID: 30242250; PMCID: PMC6154957.

[12] Liu W, Wang M, Shen L, Zhu Y, Ma H, Liu B, Ouyang L, Guo W, Xu Q, Sun Y. SHP2-mediated mitophagy boosted by lovastatin in neuronal cells alleviates parkinsonism in mice. Signal Transduct Target Ther. 2021 Jan 29;6(1):34. doi: 10.1038/s41392-021-00474-x. PMID: 33514686; PMCID: PMC7846847.

[13] Sticherling M. Psoriasis and autoimmunity. Autoimmun Rev. 2016 Dec;15(12):1167–1170. doi: 10.1016/j.autrev.2016.09.004. Epub 2016 Sep 15. PMID: 27639838.

[14] Hu SC, Yu HS, Yen FL, Lin CL, Chen GS, Lan CC. Neutrophil extracellular trap formation is increased in psoriasis and induces human β-defensin-2 production in epidermal keratinocytes. Sci Rep. 2016 Aug 5;6:31119. doi: 10.1038/srep31119. PMID: 27493143; PMCID: PMC4974609.

[15] Kim DJ, Tremblay ML, Digiovanni J. Protein tyrosine phosphatases, TC-PTP, SHP1, and SHP2, cooperate in rapid dephosphorylation of Stat3 in keratinocytes following UVB irradiation. PLoS One. 2010 Apr 22;5(4):e10290. doi: 10.1371/journal.pone.0010290. PMID: 20421975; PMCID: PMC2858656.

[16] Zhang RY, Yu ZH, Zeng L, Zhang S, Bai Y, Miao J, Chen L, Xie J, Zhang ZY. SHP2 phosphatase as a novel therapeutic target for melanoma treatment. Oncotarget. 2016 Nov 8;7(45):73817–73829. doi: 10.18632/oncotarget.12074. PMID: 27650545; PMCID: PMC5342016.

[17] Zehender A, Huang J, Györfi AH, Matei AE, Trinh-Minh T, Xu X, Li YN, Chen CW, Lin J, Dees C, Beyer C, Gelse K, Zhang ZY, Bergmann C, Ramming A, Birchmeier W, Distler O, Schett G, Distler JHW. The tyrosine phosphatase SHP2 controls TGFβ-induced STAT3 signaling to regulate fibroblast activation and fibrosis. Nat Commun. 2018 Aug 14;9(1):3259. doi: 10.1038/s41467-018-05768-3. PMID: 30108215; PMCID: PMC6092362.

[18] Li XJ, Goodwin CB, Nabinger SC, Richine BM, Yang Z, Hanenberg H, Ohnishi H, Matozaki T, Feng GS, Chan RJ. Protein-tyrosine phosphatase Shp2 positively regulates macrophage oxidative burst. J Biol Chem. 2015 Feb 13;290(7):3894–909. doi: 10.1074/jbc.M114.614057. Epub 2014 Dec 23. PMID: 25538234; PMCID: PMC4326800.

[19] Xiao P, Zhang H, Zhang Y, Zheng M, Liu R, Zhao Y, Zhang X, Cheng H, Cao Q, Ke Y. Phosphatase Shp2 exacerbates intestinal inflammation by disrupting macrophage responsiveness to interleukin-10. J Exp Med. 2019 Feb 4;216(2):337–349. doi: 10.1084/jem.20181198. Epub 2019 Jan 4. PMID: 30610104; PMCID: PMC6363431.

[20] Zhang Y, Liu H, Yao J, Huang Y, Qin S, Sun Z, Xu Y, Wan S, Cheng H, Li C, Zhang X, Ke Y. Manipulating the air-filled zebrafish swim bladder as a neutrophilic inflammation model for acute lung injury. Cell Death Dis. 2016 Nov 10;7(11):e2470. doi: 10.1038/cddis.2016.365. PMID: 27831560; PMCID: PMC5260887.

[21] Muraro SP, De Souza GF, Gallo SW, Da Silva BK, De Oliveira SD, Vinolo MAR, Saraiva EM, Porto BN. Respiratory Syncytial Virus induces the classical ROS-dependent NETosis through PAD-4 and necroptosis pathways activation. Sci Rep. 2018 Sep 21;8(1):14166. doi: 10.1038/s41598-018-32576-y. PMID: 30242250; PMCID: PMC6154957.

