## Supplementary figures and images for "SHP2 promotes NETosis through ERK5 pathway and mediates the development of psoriasis"

### FIG-S1.tif

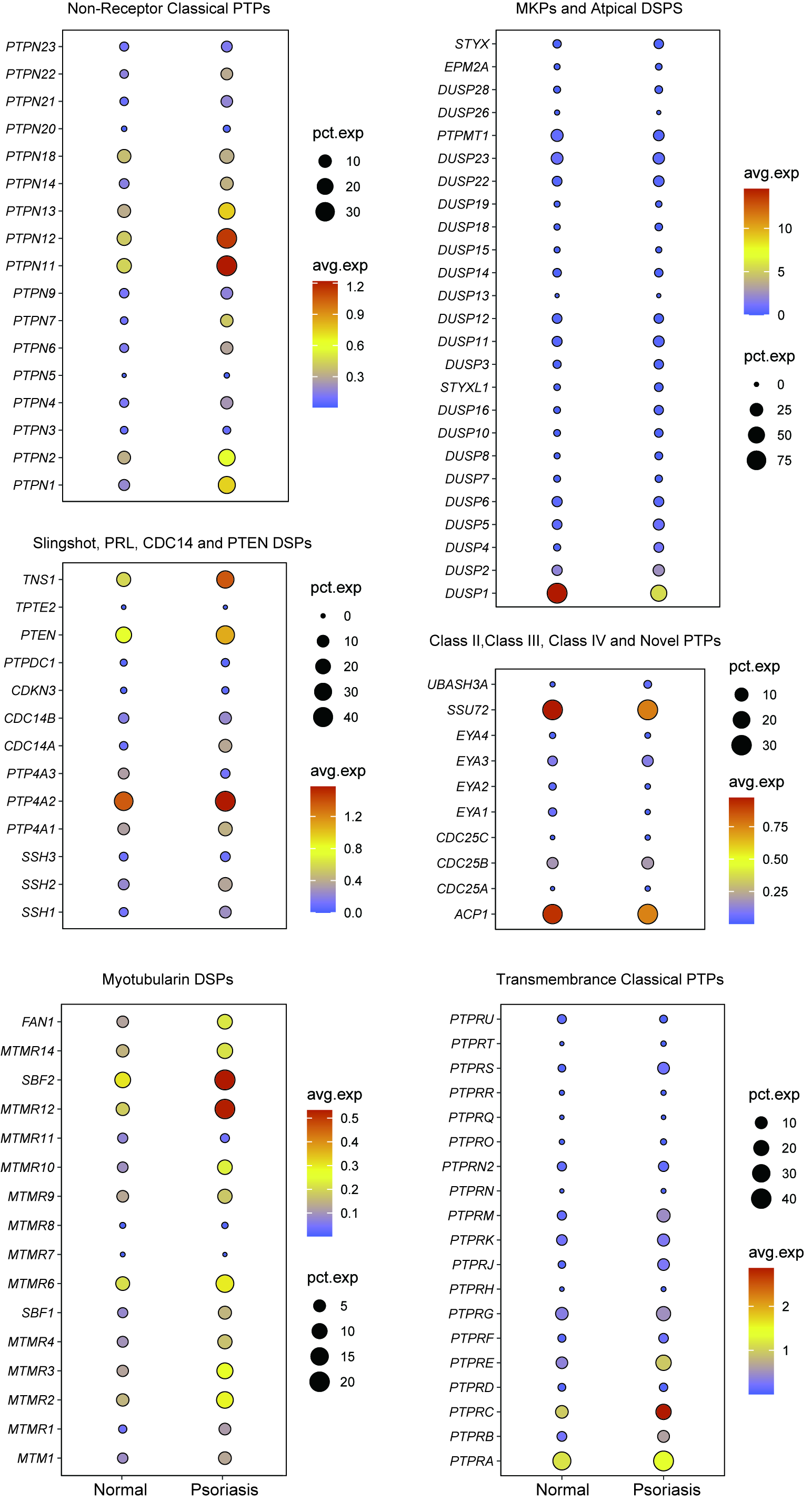

### FIG-S2.tif

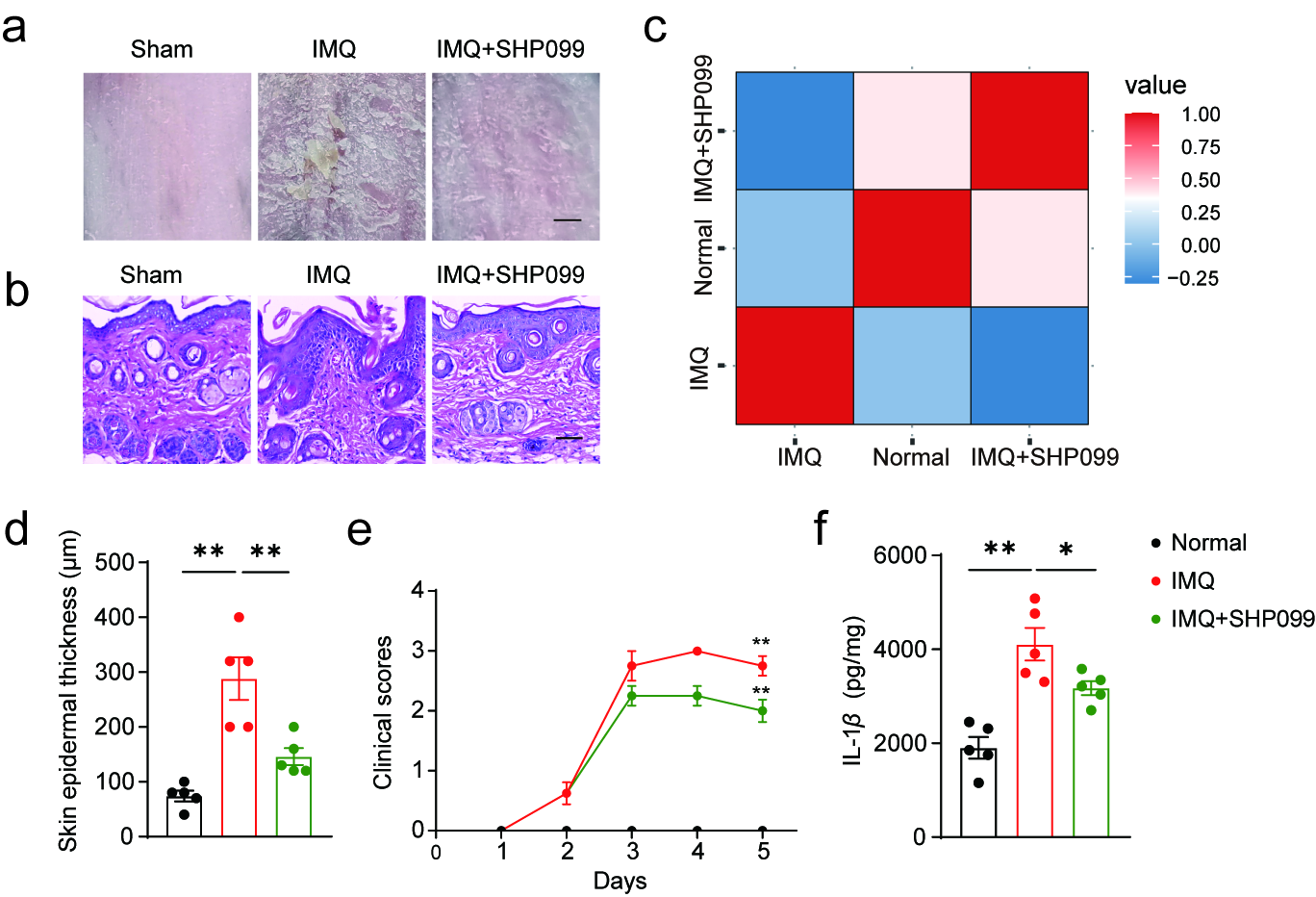

### FIG-S3.tif

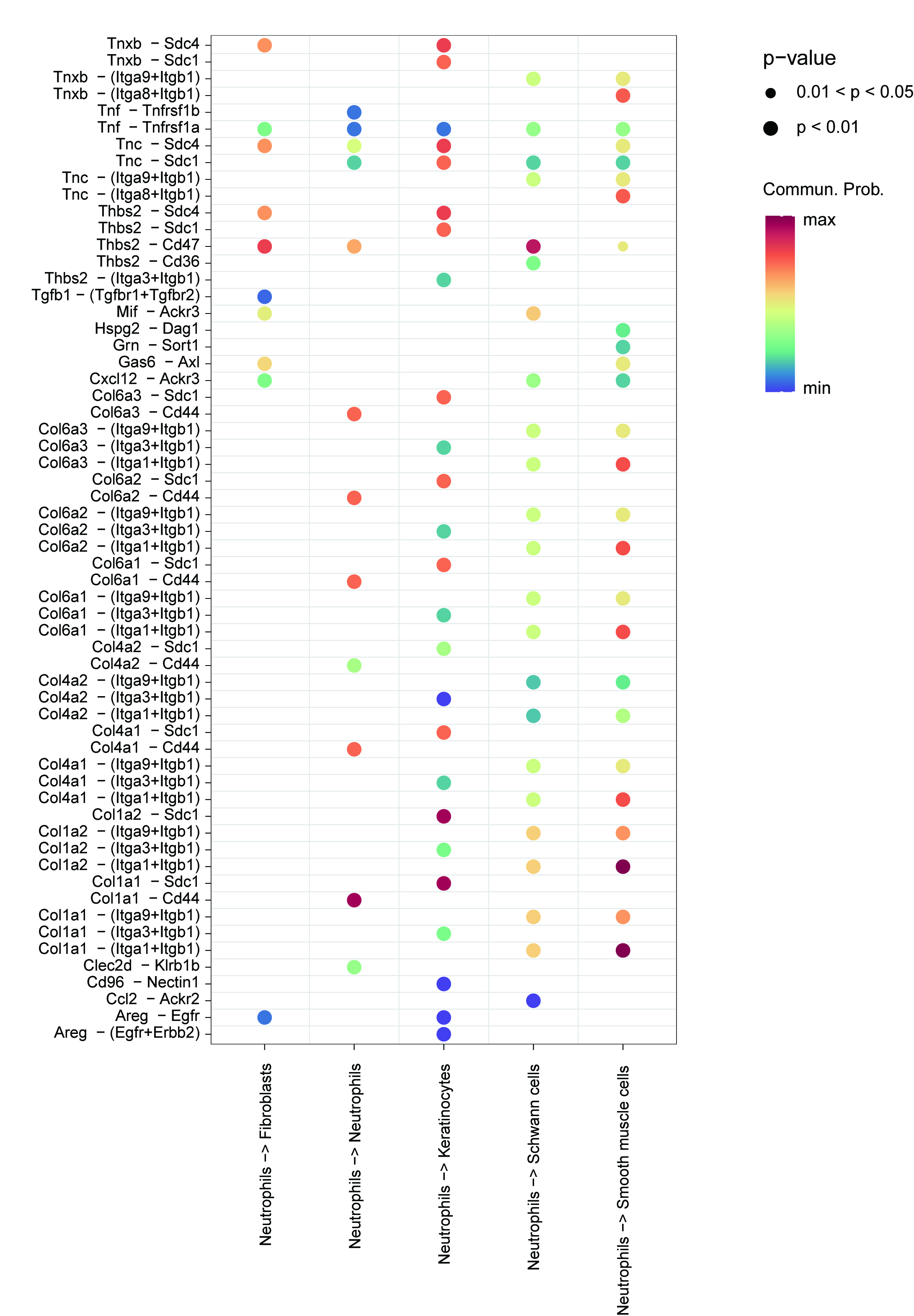
